# Cost-effectiveness of short, oral treatment regimens for rifampicin resistant tuberculosis

**DOI:** 10.1101/2022.11.08.22282060

**Authors:** Sedona Sweeney, Catherine Berry, Emil Kazounis, Ilaria Motta, Anna Vassall, Matthew Dodd, Katherine Fielding, Bern-Thomas Nyang’wa

**Affiliations:** Faculty of Public Health and Policy, London School of Hygiene & Tropical Medicine, London UK; Public Health Department OCA, Médecins Sans Frontières, UK; Faculty of Epidemiology and Population Health, London School of Hygiene & Tropical Medicine, London UK; Public Health Department OCA, Médecins Sans Frontières, the Netherlands; Faculty of Infectious and Tropical Diseases, London School of Hygiene & Tropical Medicine, London UK

## Abstract

**Introduction:** Current options for treating tuberculosis (TB) that is resistant to rifampicin (RR-TB) are few, and regimens are often long and poorly tolerated. Following recent evidence from the TB-PRACTECAL trial countries are considering programmatic uptake of 6-month, all-oral treatment regimens.

**Methods:** We used a Markov model to estimate the incremental cost-effectiveness of three regimens containing bedaquiline, pretomanid and linezolid (BPaL) with and without moxifloxacin (BPaLM) or clofazimine (BPaLC) compared with the current mix of long and short standard of care (SOC) regimens to treat RR-TB from the provider perspective in India, Georgia, Philippines, and South Africa. We estimated total costs (2019 USD) and disability-adjusted life years (DALYs) over a 20-year time horizon. Costs and DALYs were discounted at 3% in the base case. Parameter uncertainty was tested with univariate and probabilistic sensitivity analysis.

**Results:** We found that all three regimens would improve health outcomes and reduce costs compared with the current programmatic mix of long and short SOC regimens in all four countries. BPaL was the most cost-saving regimen in all countries, saving $112-$1,173 per person. BPaLM was the preferred regimen at a willingness to pay per DALY of 0.5 GDP per capita in all settings.

**Conclusions:** Our findings indicate BPaL-based regimens are likely to be cost-saving and more effective than the current standard of care in a range of settings. Countries should consider programmatic uptake of BPaL-based regimens.

## Introduction

Tuberculosis (TB) is the world’s second deadliest infection after COVID (1), with an estimated global 9.8 million people developing active TB disease and 1.5 million deaths in 2020 alone (2). Each year about 470,000 people fall ill with a strain of TB that is resistant to rifampicin (RR-TB), one of the core drugs in the standard first-line treatment regimen (2). Treatment options for RR-TB, also potentially including resistance to isoniazid (i.e. multi-drug resistant TB (MDR-TB)) and/or fluoroquinolones (pre-XDR TB), are few and despite recent advances still involve long treatment durations, high pill counts, and injections (3). Outcomes for RR-TB treatment are also poor; in 2018 the global treatment success rate for RR-TB was only 59% (2).

There have been substantial recent changes in the landscape of treatment options available for people with RR-TB. BPaL regimens, which contain bedaquiline, pretomanid, and linezolid, are all-oral and only 6 months in duration, as compared with 9-month shorter regimens and 20-month longer regimens currently used in most countries. Pretomanid as part of BPaL regimens has been approved by stringent regulatory authorities (US Federal Drug Administration and European Medicines Agency) and in another 28 countries, and prequalified by WHO (4). BPaL regimens were recommended for use under operational research conditions by WHO in 2021 (5) following evidence from an open-label, single-arm study (Nix-TB) that BPaL led to favourable outcomes 6 months after treatment completion in populations with highly drug-resistant TB (including treatment intolerant MDR-TB and pre-XDR TB), although some linezolid-related adverse events were noted (6). Subsequently, the ZeNix trial evaluated BPaL regimens with linezolid dosed at 1200 and 600mg for varying durations, and found maintenance of treatment success among the same population, with reduction of linezolid-associated adverse events at 600mg daily (7).

The TB-PRACTECAL trial has added new evidence to support the use of shortened all-oral regimens. TB-PRACTECAL is the first multi-country, randomised, controlled, open label, phase II/III trial to evaluate the safety and efficacy of drug regimens containing bedaquiline and pretomanid for the treatment of patients with pulmonary RR-TB. Trial recruitment was terminated early, and all three investigative trial arms were found to perform significantly better than the control (8). TB-PRACTECAL adds a large sample of patients representing rifampicin resistance, including patients with and without fluoroquinolone resistant tuberculosis and HIV coinfection.

Following these scientific successes and hopes for substantially reduced burden on both the health sector and TB patients through shortened treatment periods, the WHO has announced forthcoming recommendations for programmatic use of BPaL-based regimens and countries are starting to consider the potential advantages and costs of replacing the current standard of care for RR-TB (9). A cost-effectiveness analysis of the TB-PRACTECAL intervention, as implemented by Médecins Sans Frontières (MSF) in South Africa, Belarus, and Uzbekistan, is planned and cost data collection is underway (10). However, decision makers also need information on the potential ‘real world’ economic value of these newer regimens along with their clinical efficacy when planning for introduction at the programmatic level. The aim of this analysis was to estimate the likely incremental cost-effectiveness of the three TB-PRACTECAL regimens at a programmatic level, as compared with existing WHO-recommended standard of care regimens, in a variety of settings.

## Methods

We conducted a Markov model-based analysis to estimate the incremental cost-effectiveness of the three BPaL-based regimens investigated in the TB-PRACTECAL trial (described in Supplementary Table 1) for treatment of TB that is at least resistant to rifampicin, as compared with the current standard of care (SOC) in Philippines, South Africa, Georgia, and India (Table 1).

**Table 1.**
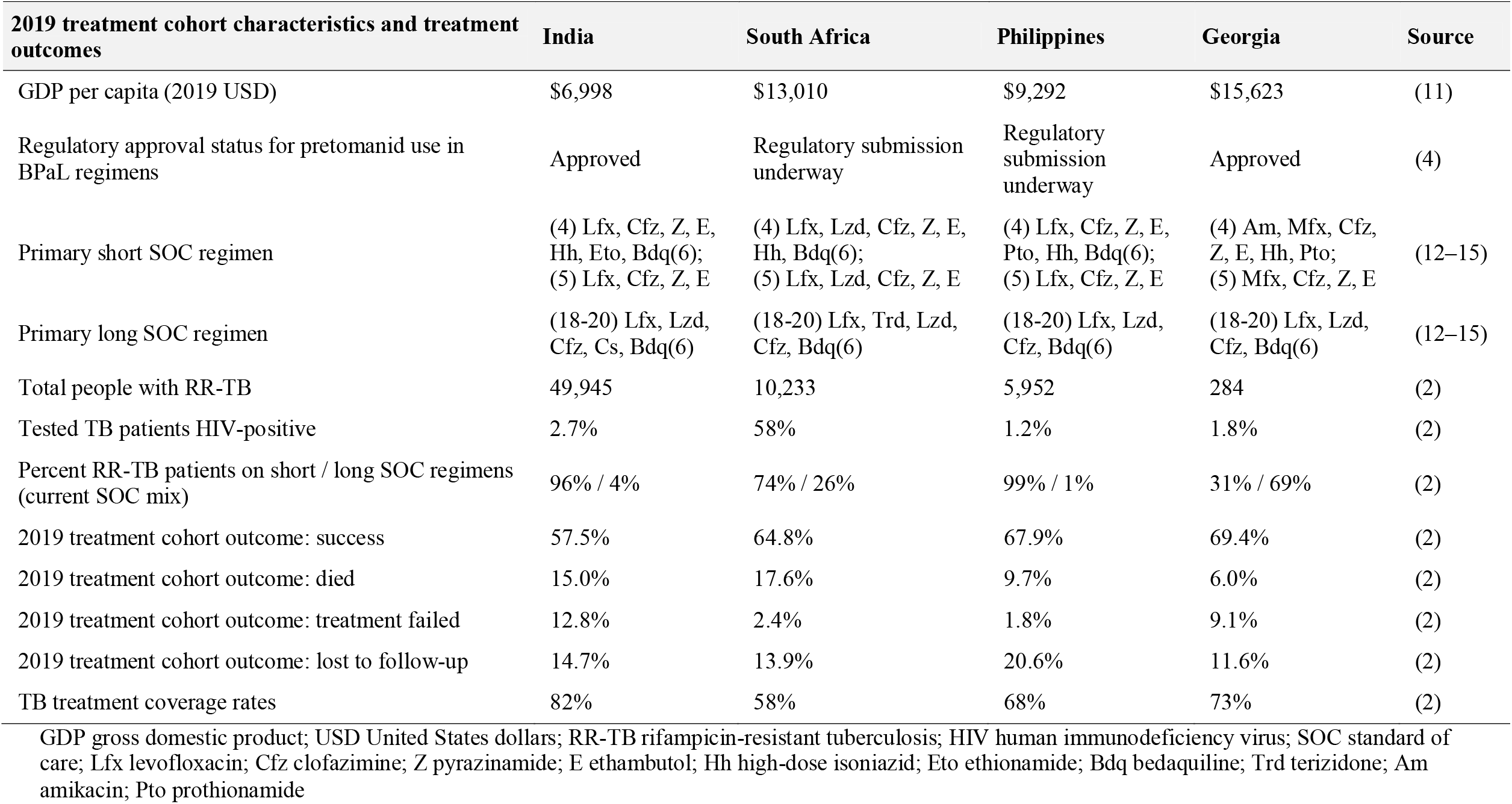
Country characteristics.

| **2019 treatment cohort characteristics and treatment outcomes** | **India** | **South Africa** | **Philippines** | **Georgia** | **Source** |
| --- | --- | --- | --- | --- | --- |
| GDP per capita (2019 USD) | \$6,998 | \$13,010 | \$9,292 | \$15,623 | (11) |
| Regulatory approval status for pretomanid use in BPaL regimens | Approved | Regulatory submission underway | Regulatory submission underway | Approved | (4) |
| Primary short SOC regimen | (4) Lfx, Cfz, Z, E, Hh, Eto, Bdq(6);<br>(5) Lfx, Cfz, Z, E | (4) Lfx, Lzd, Cfz, Z, E, Hh, Bdq(6);<br>(5) Lfx, Lzd, Cfz, Z, E | (4) Lfx, Cfz, Z, E, Pto, Hh, Bdq(6);<br>(5) Lfx, Cfz, Z, E | (4) Am, Mfx, Cfz, Z, E, Hh, Pto;<br>(5) Mfx, Cfz, Z, E | (12–15) |
| Primary long SOC regimen | (18-20) Lfx, Lzd, Cfz, Cs, Bdq(6) | (18-20) Lfx, Trd, Lzd, Cfz, Bdq(6) | (18-20) Lfx, Lzd, Cfz, Bdq(6) | (18-20) Lfx, Lzd, Cfz, Bdq(6) | (12–15) |
| Total people with RR-TB | 49,945 | 10,233 | 5,952 | 284 | (2) |
| Tested TB patients HIV-positive | 2.7% | 58% | 1.2% | 1.8% | (2) |
| Percent RR-TB patients on short / long SOC regimens (current SOC mix) | 96% / 4% | 74% / 26% | 99% / 1% | 31% / 69% | (2) |
| 2019 treatment cohort outcome: success | 57.5% | 64.8% | 67.9% | 69.4% | (2) |
| 2019 treatment cohort outcome: died | 15.0% | 17.6% | 9.7% | 6.0% | (2) |
| 2019 treatment cohort outcome: treatment failed | 12.8% | 2.4% | 1.8% | 9.1% | (2) |
| 2019 treatment cohort outcome: lost to follow-up | 14.7% | 13.9% | 20.6% | 11.6% | (2) |
| TB treatment coverage rates | 82% | 58% | 68% | 73% | (2) |

GDP gross domestic product; USD United States dollars; RR-TB rifampicin-resistant tuberculosis; HIV human immunodeficiency virus; SOC standard of care; Lfx levofloxacin; Cfz clofazimine; Z pyrazinamide; E ethambutol; Hh high-dose isoniazid; Eto ethionamide; Bdq bedaquiline; Trd terizidone; Am amikacin; Pto prothionamide

Incremental costs from the provider perspective and disability-adjusted life years (DALYs) averted were modelled over a 20-year time horizon. This model-based analysis was conducted to inform growing policy questions following the early termination of the TB-PRACTECAL trial and WHO recommendations for programmatic uptake of BPaL-based regimens (9). Methods for the economic evaluation were adapted from the planned per-protocol economic analysis (10).

### Patient and Public Involvement

The research primarily targeted policy makers, and the study question was driven by the Resource Considerations requirement in the WHO guidelines development process. An economic evaluation is considered critical to the development of public health guidelines and this research study fulfilled this need. Patients were not involved in the design of this modelling study, however the aspect of this study that involves clinical outcomes was designed and implemented with extensive engagement of communities and patients (16). Results of the study will be shared in a lay summary format with community advisory boards and distributed in clinics where the study took place. Compensation for participation to the main trial was defined in close collaboration and approval from the local ethics committees.

### Study population and settings

We developed models to reflect programmatic uptake of each regimen among all patients with RR-TB in four countries with a range of characteristics potentially relevant to global decision-making context, including burden of MDR/RR-TB, burden of HIV among people with TB, and current mix of long vs. short SOC regimens for MDR-TB. Table 1 shows relevant characteristics for each country.

### Interventions and comparators

The TB-PRACTECAL trial evaluated three six-month treatment regimens; arm 3 included bedaquiline, pretomanid and tapered-dose linezolid (600mg daily for 16 weeks then 300mg daily for the remaining 8 weeks) (BPaL). Arm 1 included the BPaL backbone with addition of moxifloxacin (400mg) (BPaLM); arm 2 included the BPaL backbone with addition of clofazimine (50-100mg) (BPaLC). Each TB-PRACTECAL regimen was compared with the current mix of SOC regimens, which was estimated from the reported number of patients enrolled on short vs. long MDR-TB regimens in data underlying the most recent Global TB Report (2). Specifications for short and long SOC regimens were taken from the national MDR treatment guidelines for each country (12–15).

### Model description

Our Markov model was structured to represent progression of a cohort of patients through different health states and activities under each treatment option (Supplementary Figure 1). Patients were assumed to enter the model with active RR-TB at the point of treatment initiation. At model entry, mean age was assumed to be 35, the average age of PRACTECAL participants (17) (Supplementary Table 2). At the end of each month, patients could transition to the next month of treatment, loss to follow-up, treatment failure, or death.

Treatment duration depended on the treatment regimen; we assumed the standardized BPaL-containing regimens lasted 24 weeks in line with the TB-PRACTECAL protocol, the standard short SOC regimen lasted 36 weeks and long SOC regimens lasted 80 weeks. Patients who were lost to follow-up (LTFU) before treatment completion had a chance of re-entering treatment each month in the first two years from the start of treatment. Following failure from the initial treatment regimen, we assumed all patients were put on a long SOC ‘rescue’ regimen regardless of their initial treatment regimen. Patients who had treatment failure on the rescue regimen moved to an ‘end of life’ state. Patients successfully completing treatment had a chance of experiencing relapse in the first four years following treatment completion; reinfection was not included in the model. Patients who relapsed had a chance of re-entering treatment following national TB treatment coverage rates. We did not model any potential reduction of onward transmission of TB due to a shorter treatment period. Key model parameters are listed in Supplementary Table 2.

### Treatment Outcomes

Outcomes for SOC regimens in the model were based on national treatment outcomes from the 2019 treatment cohort in each country, as reported to WHO (2). We assumed that there was no significant difference in treatment failure or death between standardized long and short SOC regimens (18). Following Walker et al. (19) we assumed variable risk of loss to follow-up according to the time on treatment, and adjusted treatment success rates to account for higher loss to follow-up in longer SOC regimens.

Treatment success for TB-PRACTECAL regimens were estimated using country-specific SOC treatment success, multiplied by the ratio of the probability of treatment success at 72 weeks in the interventional arms of the TB-PRACTECAL trial as compared with the SOC arm (Supplementary Table 2). Outcomes from the per-protocol population were used as this was the most conservative approach. Treatment success in the model was defined as having no unfavourable outcome (including loss to follow-up, treatment failure, recurrence, or death) at 72 weeks. Although the trial outcomes showed reduced deaths in the investigational regimens, we conservatively assumed no impact on deaths, treatment failures, or recurrence, and instead assumed improvements in treatment success resulted from reductions in loss to follow-up. This conservative approach was taken as the very low number of failures and deaths in both interventional and SOC populations observed in the TB-PRACTECAL trial may not be achievable in programmatic settings, and is tested in univariate sensitivity analysis.

Monthly transition probabilities for all outcomes were calculated using the average treatment duration for each regimen. Patient costs and outcomes were tallied over a lifetime horizon of 20 years, with cycles of one month. We chose 20 years as this was adequate to capture all relevant costs and outcomes.

The measure of effectiveness for our incremental cost-effectiveness estimates was in terms of DALYs averted. DALY weights for each health state were sourced from the most recent Global Burden of Disease study, and are reported in Supplementary Table 3 (20). Following evidence that there is a lifelong post-TB health burden even in patients with treatment success, we used a DALY weight of 0.053 for the ‘post-TB’ state (21). We used expert opinion to map adverse event symptoms to equivalent health states where adverse events were not specified in the Global Burden of Disease database. We used a multiplicative model to generate a combined disability weight for patients with adverse events (22). All DALYs were discounted at 3% in the base case (23).

### Costs

Treatment costs were estimated from a providers’ perspective using a combination of country MDR treatment guidelines, previously published estimates of prices and quantities of different services, and expert opinion (Supplementary Table 4). A provider perspective was chosen due to a lack of high-quality cost data from the patient perspective for the included countries. For Georgia, India, and Philippines, we derived the unit costs of outpatient visits, inpatient bed-days, community and other visits, and laboratory tests from the Value TB dataset, the largest multi-country TB costing study to date (24). Unit costs for health care services in South Africa, and for procedures, medications and fluids for adverse events, were sourced from the literature (25,26). All costs are reported in 2019 USD. Costs reported in USD from previous years were inflated using the United States gross domestic product (GDP) deflator (11).

We estimated the quantities of outpatient visits, inpatient bed-days, community- and other visits, and lab tests per month using the Value TB dataset, national country guidelines, and validated these through expert opinion (Supplementary Figure 2) (12,15,24). Where data were available, we used the ‘real world’ quantities of a wide range of services, as delivered in health facilities observed in the Value TB study. This often differed from country guidelines but was a better representation of total costs incurred during treatment. Quantities were estimated separately for intensive and continuation phases for all SOC regimens, including a two-month inpatient treatment period for all regimens in Georgia. Quantities of services per month for investigational regimens were assumed to follow the pattern of the first six months of care for the short SOC regimen. Expert opinion was used to estimate the quantities of outpatient visits and inpatient bed-days per average patient for different adverse events. Patients who were lost to follow-up (LTFU) were assumed to incur costs of one LTFU tracing call or home visit per month until death or return to care.

Drug prices were sourced from the Global Drug Facility Catalogue for all countries (27). Drug quantities were modelled according to each country’s recommended SOC treatment regimen using standard recommended dosage for an adult weighing 51-70kg. All costs were discounted at a rate of 3% in the base case (23).

### Uncertainty and sensitivity analyses

Incremental costs and effects were estimated using mean parameter values in the base case. Parameter uncertainty was assessed in a probabilistic sensitivity analysis, varying all parameters over 200 simulations. Risk ratios for treatment success were varied following a log normal distribution. Unit costs were varied using a gamma distribution, and all DALY weights were varied following a beta distribution. We also conducted a univariate sensitivity analysis to test model sensitivity to changes in key parameters, including non-drugs costs, prices for bedaquiline and pretomanid, likelihood of return to care after loss to follow-up, discount rate, and valuation of the post-TB health state. We estimated the incremental costs and effects of interventional regimens with each of these key parameters halved and doubled to represent extreme possible values. We also tested the base-case assumption that improvements in effectiveness from the TB-PRACTECAL regimens resulted from reductions in losses to follow-up; we ran alternative scenarios with reductions in deaths and treatment failures instead. We did not characterize heterogeneity or distributional effects as subgroup analyses were not performed on these preliminary outcomes from the trial.

## Results

Table 2 shows results from the base-case analysis. In all settings the current SOC mix is the most expensive treatment option, and BPaL is the cheapest regimen. The cost savings associated with a move from the current SOC mix to BPaL for all MDR/RR-TB patients range from $1,173 per person in South Africa to $112 per person in India. BPaLM would save $80-$904and avert 0.7-1.3 DALYs per person.

**Table 2.** Base case results.

|  |  |  | Comparison with current SOC mix |  |
| --- | --- | --- | --- | --- |
| Country and regimen | Total costs per person | Total DALYs per person | Incremental Costs Per Person | Incremental DALYs Averted Per Person |
| **Philippines** |  |  |  |  |
| Current SOC regimen mix (99% short, 1% long) | \$1,329 | 5.4 | | |
| BPaL | \$1,078 | 5.4 | -\$251 | 0.0 |
| BPaLC | \$1,174 | 5.3 | -\$155 | 0.1 |
| BPaLM | \$1,124 | 4.6 | -\$204 | 0.8 |
| **South Africa** |  |  |  |  |
| Current SOC regimen mix (74% short, 26% long) | \$4,517 | 6.8 | | |
| BPaL | \$3,344 | 6.6 | -\$1,173 | 0.2 |
| BPaLC | \$3,470 | 6.5 | -\$1,047 | 0.3 |
| BPaLM | \$3,520 | 6.0 | -\$997 | 0.8 |
| **India** |  |  |  |  |
| Current SOC regimen mix (96% short, 4% long) | \$978 | 6.4 | | |
| BPaL | \$867 | 6.4 | -\$112 | 0.0 |
| BPaLC | \$952 | 6.3 | -\$27 | 0.1 |
| BPaLM | \$899 | 5.7 | -\$80 | 0.7 |
| **Georgia** |  |  |  |  |
| Current SOC regimen mix<br>(31% short, 69% long) | \$4,211 | 4.7 | | |
| BPaL | \$3,228 | 4.3 | -\$983 | 0.4 |
| BPaLC | \$3,327 | 4.2 | -\$883 | 0.6 |
| BPaLM | \$3,307 | 3.5 | -\$904 | 1.3 |

SOC standard of care; DALYs disability-adjusted life years. All costs presented in 2019 US Dollars.

Figure 1 shows the average lifetime costs for each cohort. Reducing the duration of treatment episodes through investigational regimens reduced non-pharmaceutical costs; this change was most substantial in South Africa and Georgia where visit costs were high. The reduction in non-pharmaceutical costs was less substantial in India and Philippines, where the costs of non-tradeable inputs such as human resources are very low, and where more patients are currently on shorter SOC regimens. Tradeable drugs costs for investigational regimens are slightly higher than those for short SOC regimens in all countries. The total costs incurred for adverse events and loss to follow-up tracing were typically higher in SOC regimens, but this did not have a substantial effect on total costs.

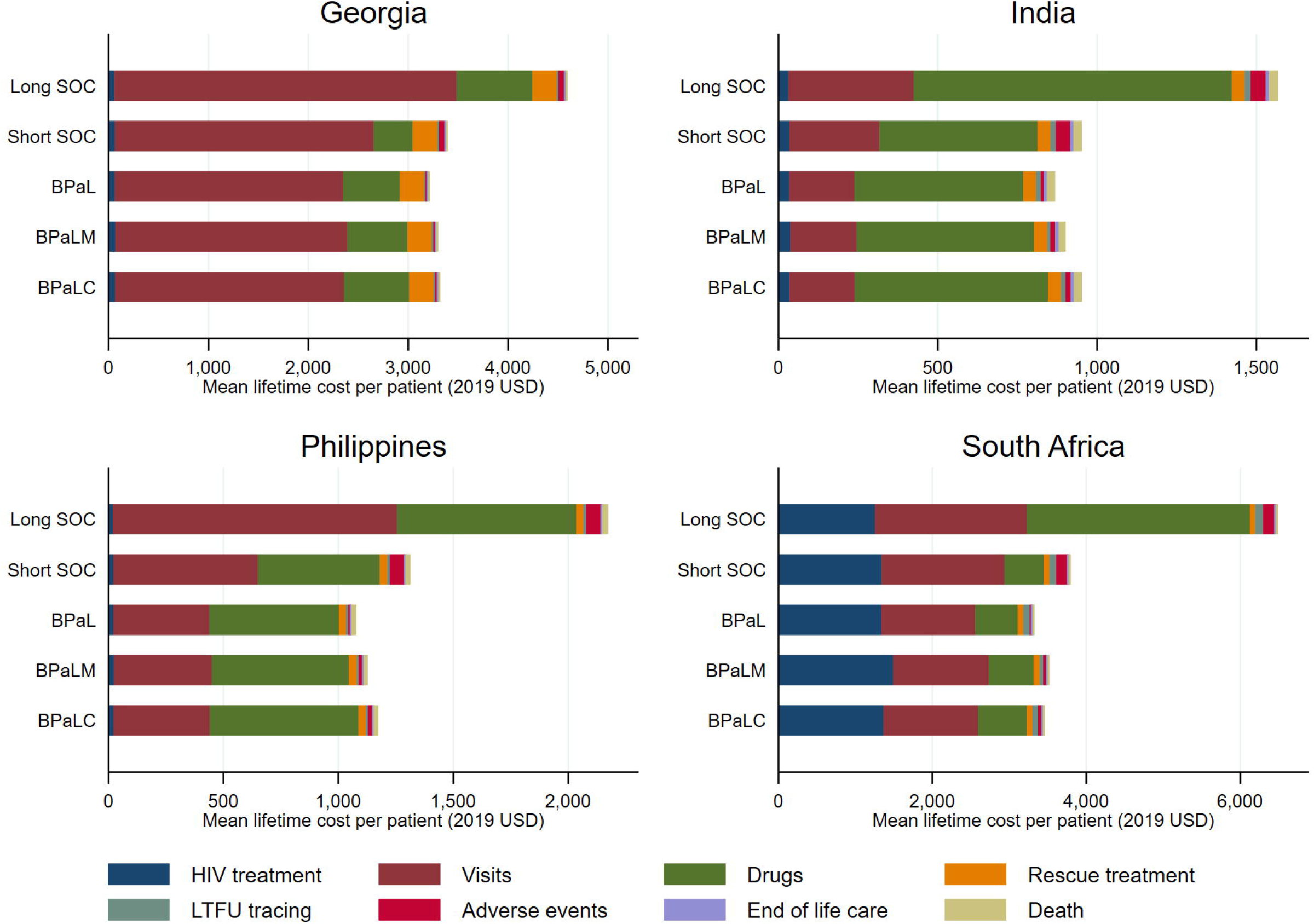

In the probabilistic sensitivity analysis, all simulations were cost saving for all three investigational regimens in Philippines, South Africa, and Georgia (Figure 2, Supplementary Table 5). In India most simulations for all regimens were cost saving (99% BPaL, 74% BPaLC, and 94% BPaLM simulations). In all countries, the mean DALYs averted across 200 simulations for all regimens were positive, however there were high levels of uncertainty around this outcome in Philippines and India.

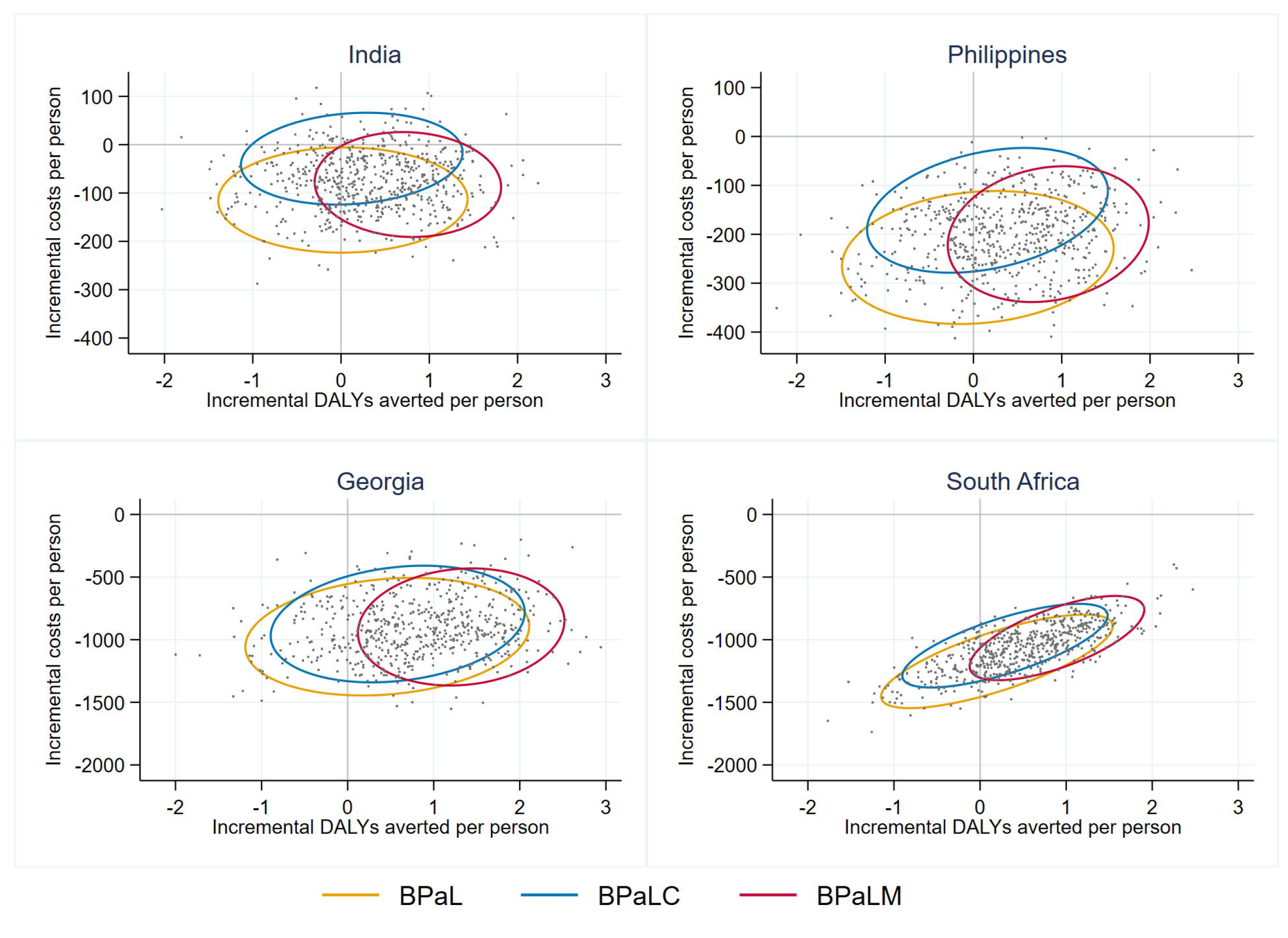

Our univariate sensitivity analysis showed the model was most sensitive to non-pharmaceutical costs in Georgia and South Africa where visit costs were high, and most sensitive to drugs costs in India and Philippines where visit costs were low (Supplementary Figure 2). Only in India, where the incremental cost savings associated with investigational regimens were lowest, did a change in parameters lead to a slight positive incremental cost of investigational regimens as compared with the current mix. Health outcomes in the model were sensitive to the choice of discount rate. Changing assumptions around the post-TB DALY weight and likelihood of return from LTFU did not have a substantial impact on findings. Figure 3 shows cost-effectiveness acceptability curves for the three investigational regimens and current SOC mix in each country. At a willingness-to-pay per DALY averted of 0.5 GDP per capita, BPaLM is the preferred regimen in all countries.

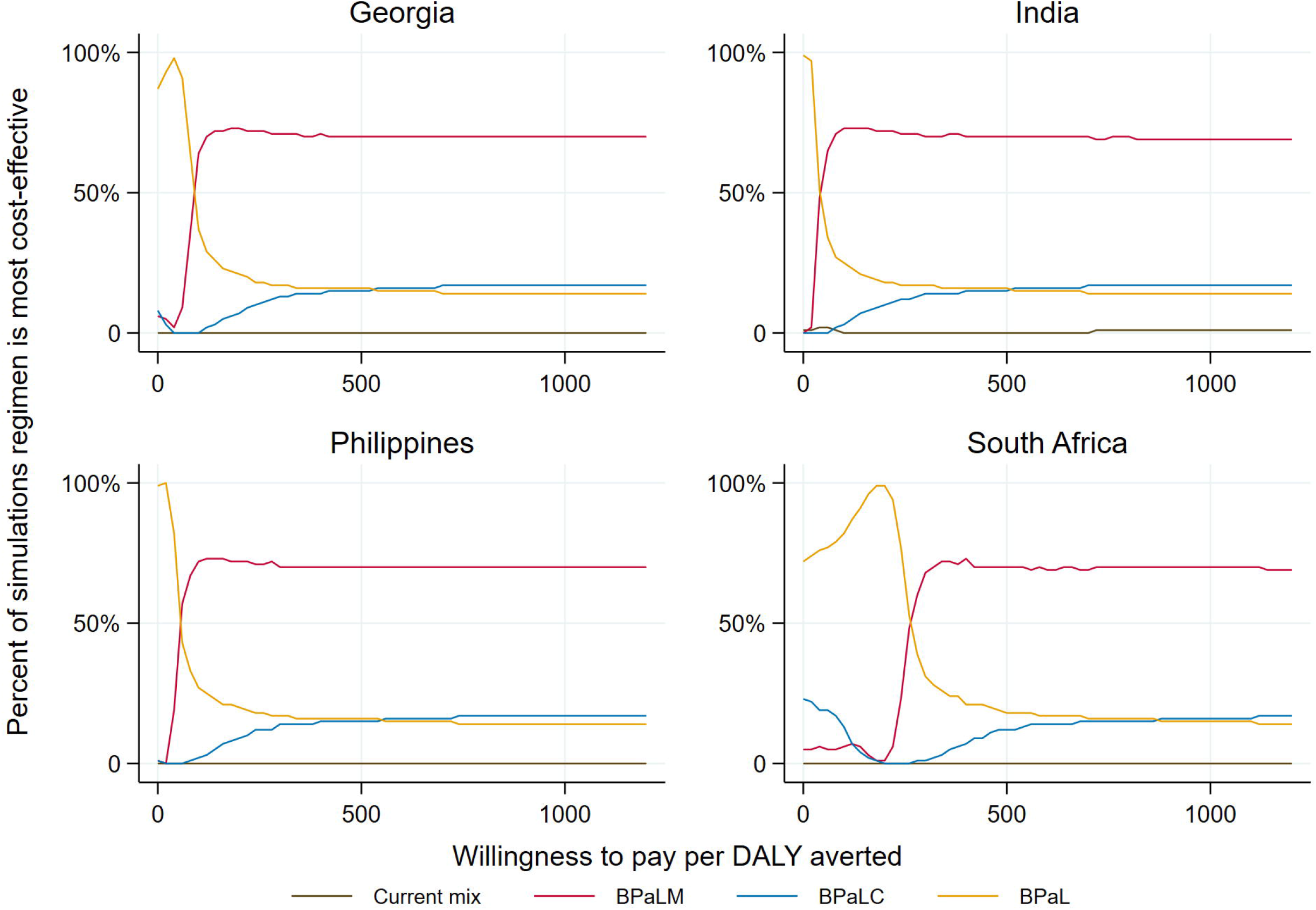

## Discussion

We estimated the incremental cost-effectiveness of implementing BPaLM, BPaLC and BPaL regimens on a programmatic level, as compared with the current SOC mix in Philippines, South Africa, Georgia and India using a model-based analysis. We found that all three regimens were highly likely to be cost saving in all settings. Our base-case estimates of the incremental effect of investigational regimens were highly conservative and assumed no change in deaths, treatment failures, or relapse from the SOC.

This is the first paper to evaluate the cost-effectiveness of replacing the current standard of care with BPaL-based regimens for RR-TB patients with and without fluoroquinolone resistance. Two previously published modelling analyses evaluated the cost-effectiveness and budgetary impact of BPaL for patients with pre-XDR TB and treatment-intolerant or non-responsive MDR-TB using data from the Nix trial (28,29). In South Africa, Georgia and Philippines, BPaL was found to be cost-saving for this population; these results were sensitive to drugs prices and assumptions around loss to follow-up (29). In Indonesia, Kyrgyzstan, and Nigeria, adoption of the BPaL regimen was estimated to lead to a reduction in XDR-related spending between 15-32% of current expenditure (28). Our findings support these broad conclusions that BPaL-based regimens are cost-saving and adds evidence for patients who have TB strains that are resistant rifampicin but susceptible to fluoroquinolones.

The main driver of the cost savings observed in our model was a reduction of non-drugs costs through reduced time in treatment. Whether these potential cost savings can be realised depends to some extent on the ability of the health system to shift existing capacity to different health services. Reduced numbers of visits for TB patients may, for example, free up clinician or nurse time that could instead be used to deliver other services. If time or other resources cannot be repurposed, and efficiency of TB services reduces through increased excess capacity, the potential cost savings in this analysis may not be fully realised.

Our analysis was restricted to the provider perspective due to data limitations. The potential savings through shortened treatment periods from the household perspective are substantial. Globally, 87% of people with drug-resistant TB face catastrophic TB-related costs (2). Reducing costs incurred at the patient- and household level by shortening the treatment period could mean saving countless households from falling into, or further into, poverty because of their TB.

We present here the results of a modelling study, which is based on several assumptions, and should be taken as an aid to decision-making rather than a statement of scientific fact. Our model has several limitations. The model did not distinguish between people with RR-TB that was resistant to or susceptible to fluoroquinolones, as TB-PRACTECAL outcomes were not estimated separately for these two patient groups. The model was unable to represent transmission impacts of shortened regimens, or evolution of TB strains that were resistant to new drugs in the BPaL-based regimens. Our model also assumed that it would be possible to fully replace current SOC regimens with BPaL-based regimens in all settings. This might not be possible due to existing resistance profiles or drug intolerances or purchasing limits for BPaL drugs. There also remains a lack of evidence in certain cohorts such as pregnant women, children under 15, and people with TB meningitis.

## Conclusions

There is now consistent evidence that 6-month bedaquiline-based regimens are likely to be cost saving at current regimen prices. Programmatic uptake of these regimens could improve treatment success rates for RR-TB and free up resources for investment in other areas of TB programmes. As countries consider shifting their current treatment strategy to shortened all-oral regimens, it is critical that TB programmes consider how best to repurpose these savings. Investment in reducing loss to follow-up through improving patient support, expanding TB case finding efforts, or improving TB prevention efforts in countries with high HIV prevalence could further country progress towards End TB targets, moving us closer to a world without TB.

## Supporting information

CHEERS checklist

## Supporting Information

Supplementary Figure 1 Markov model diagram

Supplementary Figure 2 Visit quantities per episode

Supplementary Figure 3 Univariate sensitivity analysis

Supplementary Table 1 Specification of regimens

Supplementary Table 2 Model parameters

Supplementary Table 3 DALY weights for all health states in the model

Supplementary Table 4 Unit costs

Supplementary Table 5 Probabilistic sensitivity analysis results

## Declarations

### Ethics approval

This is a model-based analysis using previously published data and did not require ethical approval.

### Availability of Data and Materials

No primary data was collected for this study. The cost-effectiveness model can be accessed on contact with the corresponding author. The cost data that was included in this cost-effectiveness model is available online at: https://dataverse.harvard.edu/dataverse/Value-TB.

### Authors Contributions

SS conceptualised the study, conducted the cost-effectiveness analysis and drafted the manuscript. CB, EK, IM, AV, MD, KF and BTN input on the methodology, critically revised the manuscript and approved the final submitted version of the manuscript. The corresponding author attests that all listed authors meet authorship criteria and that no others meeting the criteria have been omitted.

## Acknowledgements

We thank Gabriela B Gomez and Lyndon P James for their advice on study methodology.

