## Supplementary material for "Cost-effectiveness of short, oral treatment regimens for rifampicin resistant tuberculosis": CHEERS checklist

### CHEERS 2022 Checklist

| Topic | No. | Item | Location where item is reported |
| --- | --- | --- | --- |
| Title |  |  |  |
|  | 1 | Identify the study as an economic evaluation and specify the interventions being compared. | Title |
| Abstract |  |  |  |
|  | 2 | Provide a structured summary that highlights context, key methods, results, and alternative analyses. | Abstract |
| Introduction |  |  |  |
| Background and objectives | 3 | Give the context for the study, the study question, and its practical relevance for decision making in policy or practice. | Introduction section |
| Methods |  |  |  |
| Health economic analysis plan | 4 | Indicate whether a health economic analysis plan was developed and where available. | Methods section, second paragraph |
| Study population | 5 | Describe characteristics of the study population (such as age range, demographics, socioeconomic, or clinical characteristics). | Study population and settings section |
| Setting and location | 6 | Provide relevant contextual information that may influence findings. | Study population and settings section; Table 1 |
| Comparators | 7 | Describe the interventions or strategies being compared and why chosen. | Interventions and comparators section |
| Perspective | 8 | State the perspective(s) adopted by the study and why chosen. | Costs section, first paragraph |
| Time horizon | 9 | State the time horizon for the study and why appropriate. | Methods section, second paragraph |
| Discount rate | 10 | Report the discount rate(s) and reason chosen. | Treatment outcomes section, last paragraph; Costs section, last paragraph |
| Selection of outcomes | 11 | Describe what outcomes were used as the measure(s) of benefit(s) and harm(s). | Treatment outcomes section |
| Measurement of outcomes | 12 | Describe how outcomes used to capture benefit(s) and harm(s) were measured. | Treatment outcomes section |
| Valuation of outcomes | 13 | Describe the population and methods used to measure and value outcomes. | Treatment outcomes section |
| Measurement and valuation of resources and costs | 14 | Describe how costs were valued. | Costs section |
| Currency, price date, and conversion | 15 | Report the dates of the estimated resource quantities and unit costs, plus the currency and year of conversion. | Costs section, first paragraph |
| Rationale and description of model | 16 | If modelling is used, describe in detail and why used. Report if the model is publicly available and where it can be accessed. | Model description section |
| Analytics and assumptions | 17 | Describe any methods for analysing or statistically transforming data, any extrapolation methods, and approaches for validating any model used. | Uncertainty and sensitivity analysis section |
| Characterising heterogeneity | 18 | Describe any methods used for estimating how the results of the study vary for subgroups. | Uncertainty and sensitivity analysis section |
| Characterising distributional effects | 19 | Describe how impacts are distributed across different individuals or adjustments made to reflect priority populations. | Uncertainty and sensitivity analysis section |
| Characterising uncertainty | 20 | Describe methods to characterise any sources of uncertainty in the analysis. | Uncertainty and sensitivity analysis section |
| Approach to engagement with patients and others affected by the study | 21 | Describe any approaches to engage patients or service recipients, the general public, communities, or stakeholders (such as clinicians or payers) in the design of the study. | Patient and public involvement section |
| Results |  |  |  |
| Study parameters | 22 | Report all analytic inputs (such as values, ranges, references) including uncertainty or distributional assumptions. | Methods section, Table 1, Supplementary Tables 1-4 |
| Summary of main results | 23 | Report the mean values for the main categories of costs and outcomes of interest and summarise them in the most appropriate overall measure. | Results section; Table 2; Figure 1 |
| Effect of uncertainty | 24 | Describe how uncertainty about analytic judgments, inputs, or projections affect findings. Report the effect of choice of discount rate and time horizon, if applicable. | Results section; Figures 2-3; Supplementary Table 5; Supplementary Figure 3 |
| Effect of engagement with patients and others affected by the study | 25 | Report on any difference patient/service recipient, general public, community, or stakeholder involvement made to the approach or findings of the study | Not reported |
| Discussion |  |  |  |
| Study findings, limitations, generalisability, and current knowledge | 26 | Report key findings, limitations, ethical or equity considerations not captured, and how these could affect patients, policy, or practice. | Discussion section |
| Other relevant information |  |  |  |
| Source of funding | 27 | Describe how the study was funded and any role of the funder in the identification, design, conduct, and reporting of the analysis | Funding declaration |
| Conflicts of interest | 28 | Report authors conflicts of interest according to journal or International Committee of Medical Journal Editors requirements. | Competing interests declaration |

*From:* Husereau D, Drummond M, Augustovski F, et al. Consolidated Health Economic Evaluation Reporting Standards 2022 (CHEERS 2022) Explanation and Elaboration: A Report of the ISPOR CHEERS II Good Practices Task Force. Value Health 2022;25. <doi:10.1016/j.jval.2021.10.008>
