## Supplementary figures and images for "Cost-effectiveness of short, oral treatment regimens for rifampicin resistant tuberculosis"

### Supplementary Figure 1

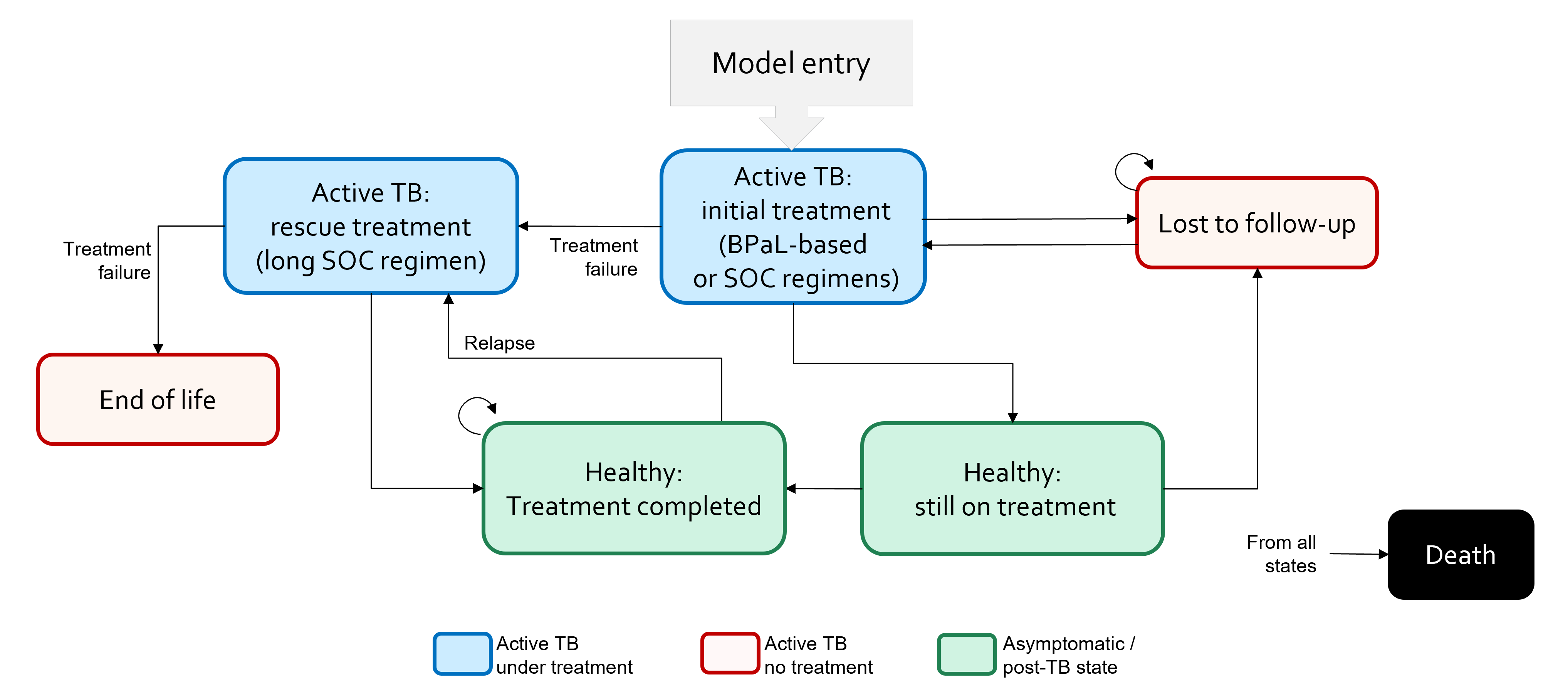

### Supplementary Figure 2

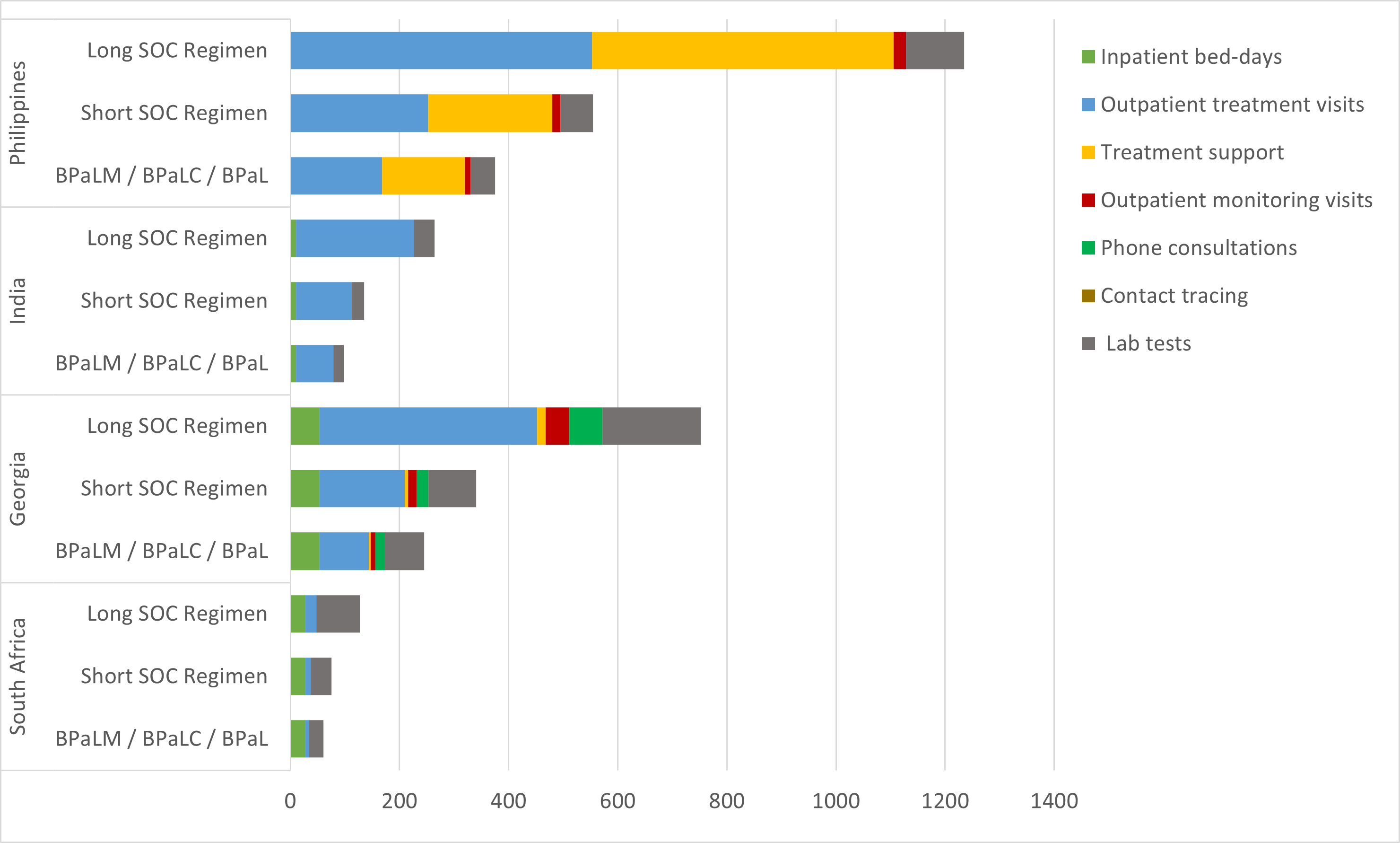

### Supplementary Figure 3

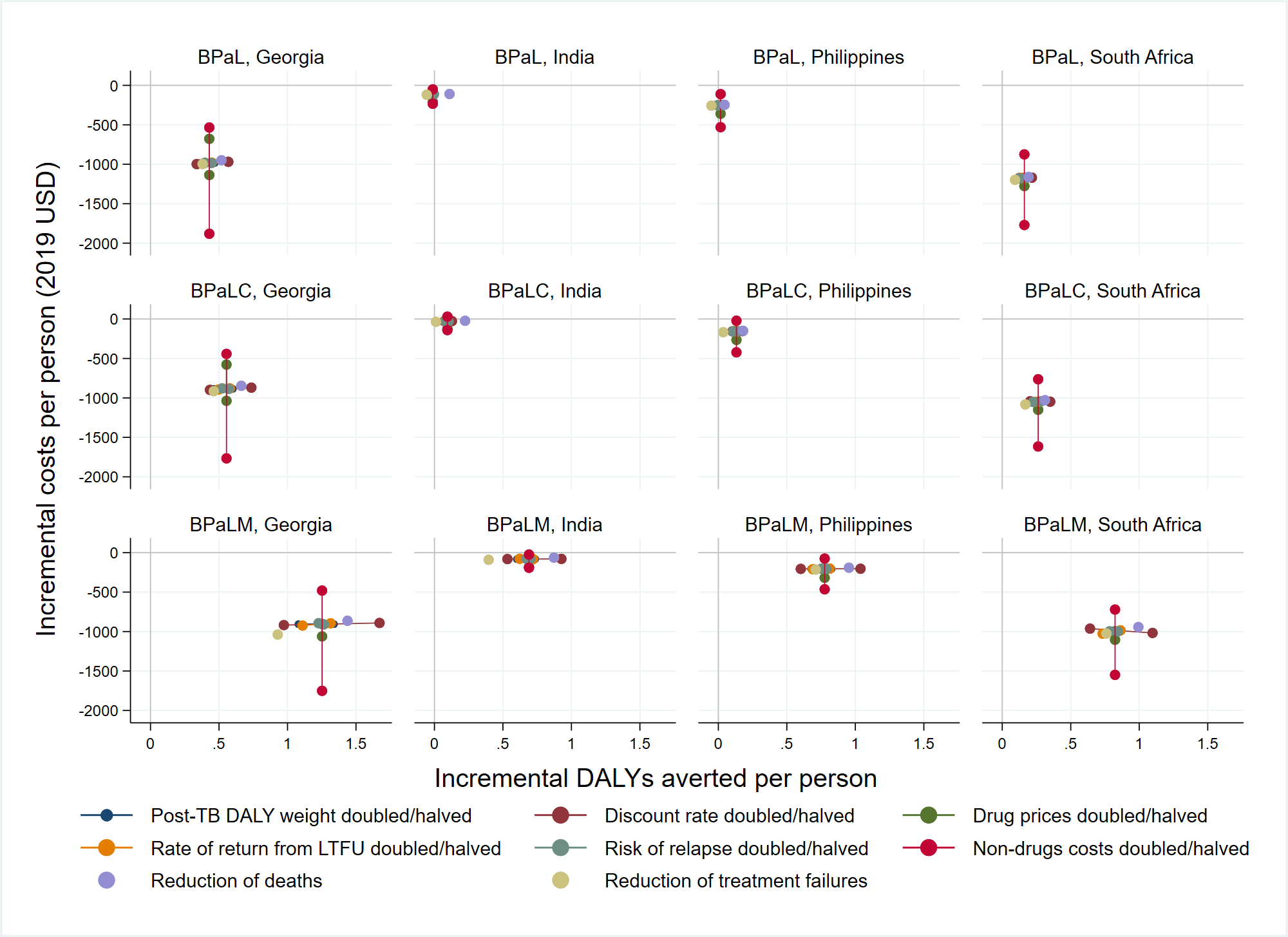
