## Supplementary Tables for "Cost-effectiveness of short, oral treatment regimens for rifampicin resistant tuberculosis"

### Supplementary Tables and Figures

###### Supplementary Table 1 Specification of regimens

| **Drug** | **BPaLM regimen** | **BPaLC regimen** | **BPaL regimen** |
| --- | --- | --- | --- |
| Bedaquiline (Bdq) | 400 mg once daily for 2 weeks followed by 200 mg 3 times per week for 22 weeks | 400 mg once daily for 2 weeks followed by 200 mg 3 times per week for 22 weeks | 400 mg once daily for 2 weeks followed by 200 mg 3 times per week for 22 weeks |
| Pretomanid (Pa) | 200mg once daily | 200mg once daily | 200mg once daily |
| Linezolid (L) | 600mg daily for 16 weeks then 300mg daily for the remaining 8 weeks or earlier when moderately tolerated | 600mg daily for 16 weeks then 300mg daily for the remaining 8 weeks or earlier when moderately tolerated | 600mg daily for 16 weeks then 300mg daily for the remaining 8 weeks or earlier when moderately tolerated |
| Moxifloxacin (Mfx) | 400 mg once daily |  |  |
| Clofazimine (Cfz) |  | 50 mg (less than 33 kg), 100 mg (more than 33 kg) |  |

###### Supplementary Table 2 Model Parameters

| **Parameter** | **Value** | **SE** | **PSA Distribution** | **Reference** |
| --- | --- | --- | --- | --- |
| Average age at model start (years) | 35 |  | N/A | assumption |
| Average body weight (kilograms) | 51-70 |  | N/A | assumption |
| Discount rates for costs and effects | 3% |  | N/A | (16) |
| Risk ratio for treatment success: BPaLM | 1.10 | 0.07 | Log normal | (17) |
| Risk ratio for treatment success: BPaLC | 1.02 | 0.08 | Log normal | (17) |
| Risk ratio for treatment success: BPaL | 1.01 | 0.08 | Log normal | (17) |
| Annual risk of relapse in year 1 post-treatment | 2.80% | 0.40% | Normal | (18) |
| Annual risk of relapse in year 2 post-treatment | 1.00% | 0.30% | Normal | (18) |
| Annual risk of relapse in year 3 post-treatment | 0.40% | 0.20% | Normal | (18) |
| Annual risk of relapse in year 4 post-treatment | 0.30% | 0.20% | Normal | (18) |
| Hazard ratio for relapse among people with HIV | 2.40 |  | N/A | (19) |
| Annual likelihood of return to care after LTFU | 28% | 3% | Normal | (20) |
| Access to end of life care | 25% | 5% | Normal | assumption |
| Monthly probability of death for end-of-life care | 6.86% | 0.69% | Beta | (21) |
| Monthly probability of death following LTFU | 6.86% | 0.69% | Beta | (21) |

SE standard error; PSA probabilistic sensitivity analysis; N/A not applicable; HIV human immunodeficiency virus; LTFU loss to follow-up

###### Supplementary Table 3 DALY weights for all health states in the model

| **Condition** | **DALY Weight** | **Health state description** | **SE** | **PSA**  **Distribution** | **Source** |
| --- | --- | --- | --- | --- | --- |
| Post-TB | 0.053 |  |  | Beta | [15] |
| HIV, no active TB | 0.125 | Has weight loss, fatigue, and frequent infections. | 0.07 | Beta | [12] |
| Active TB, no HIV | 0.333 | Has a persistent cough and fever, is short of breath, feels weak, and has lost a lot of weight. | 0.06 | Beta | [12] |
| Active TB & HIV | 0.439 | combined disability weight | 0.02 | Beta | [12] |
| End of life | 0.540 | Has lost a lot of weight and regularly uses strong medication to avoid constant pain. The person has no appetite, feels nauseous, and needs to spend most of the day in bed. | 0.09 | Beta | [12] |
| Death | 1 |  |  | Beta | [12] |
| Liver dysfunction (grade 3 and above) | 0 | Asymptomatic | - | Beta | [10, 12] |
| Pancreatitis (grade 3 and above) | 0.114 | Has pain in the belly and feels nauseous. The person has difficulties with daily activities. | 0.02 | Beta | [10, 12] |
| Anaemia (grade 3 and above) | 0.052 | Has moderate fatigue, weakness, and shortness of breath after exercise, making daily activities more difficult. | 0.01 | Beta | [10, 12] |
| Neutropenia (grade 3 and above) | 0 | Asymptomatic | - | Beta | [10, 12] |
| QTcF prolongation (grade 3 or above) | 0 | Asymptomatic | - | Beta | [10, 12] |
| Vomiting (grade 3 and above) | 0.114 | Has pain in the belly and feels nauseous. The person has difficulties with daily activities. | 0.02 | Beta | [10, 12] |
| Renal disfunction (grade 3 and above) | 0.051 | Has fever and aches, and feels weak, which causes some difficulty with daily activities. | 0.01 | Beta | [10, 12] |

DALY disability-adjusted life year; SE standard error; PSA probabilistic sensitivity analysis; QT corrected for heart rate by Fridericia's cube root formula

###### Supplementary Table 4 Unit costs

| **Service** | **Georgia** | **India** | **South Africa** | **Philippines** | **PSA**  **Distribution** | **Source** |
| --- | --- | --- | --- | --- | --- | --- |
| Outpatient diagnostic visit | $3.69 ($0.70) | $1.11 ($0.21) | $12.00 ($2.26) | $1.99 ($0.41) | Gamma | (27,28) |
| Outpatient treatment support visit | $2.42 ($0.45) | $1.44 ($0.27) | $12.00 ($2.26) | $0.42 ($0.08) | Gamma | (27,28) |
| Outpatient treatment visit | $2.13 ($0.33) | $1.15 ($0.22) | $12.00 ($2.26) | $1.68 ($0.29) | Gamma | (27,28) |
| Outpatient monitoring visit | $3.59 ($0.65) | $2.81 ($0.53) |  | $2.05 ($0.36) | Gamma | (27) |
| Inpatient bed-day | $38.28 ($3.09) | $18.06 ($3.41) | $49.03 ($3.96) | $26.63 ($2.03) | Gamma | (27,28) |
| Community-level treatment visit | $1.88 ($0.12) | $0.54 ($0.10) |  |  | Gamma | (27) |
| Community-level other visit |  | $1.70 ($0.32) |  |  | Gamma | (27) |
| Lost to follow-up tracing: phone calls | $1.24 ($0.22) |  |  | $0.70 ($0.12) | Gamma | (27) |
| Lost to follow-up tracing: home visit |  |  | $9.43 ($2.57) |  | Gamma | (28) |
| Phone consultation | $0.53 ($0.10) |  |  |  | Gamma | (27) |
| Contact tracing | $0.50 ($0.30) |  |  |  | Gamma | (27) |
| Lab tests |  |  |  |  |  |  |
| Sputum collection | $1.63 ($0.24) |  |  |  | Gamma | (27) |
| Ziehl-Neelsen smear microscopy | $5.59 ($0.84) | $1.90 ($0.36) | $6.95 ($1.04) | $2.53 ($0.39) | Gamma | (27,28) |
| Solid sputum culture | $8.56 ($1.29) | $1.90 ($0.36) |  | $23.29 ($3.17) | Gamma | (27) |
| Sputum culture | $15.55 ($2.33) |  |  |  | Gamma | (27) |
| Electrocardiogram | $1.89 ($0.26) | $0.99 ($0.19) | $14.35 ($2.15) | $3.50 ($0.63) | Gamma | (27,28) |
| HIV rapid test | $2.58 ($0.28) | $1.29 ($0.24) |  | $3.11 ($0.42) | Gamma | (27) |
| Full haemogram | $4.01 ($0.50) | $0.65 ($0.12) |  | $2.69 ($0.96) | Gamma | (27) |
| Creatinine |  | $0.59 ($0.11) |  | $1.78 ($0.27) | Gamma | (27) |
| Blood sugar | $1.01 ($0.15) | $0.78 ($0.15) |  | $2.75 ($0.59) | Gamma | (27) |
| Thyroid-stimulating hormone test |  | $2.69 ($0.51) |  |  | Gamma | (27) |
| Chest Xray (film) | $4.45 ($0.44) | $1.99 ($0.37) |  | $2.75 ($0.51) | Gamma | (27) |
| Chest Xray (digital) | $2.63 ($0.84) | $2.05 ($0.39) |  | $1.97 ($0.50) | Gamma | (27) |
| Fasting blood sugar |  |  |  | $1.55 ($0.23) | Gamma | (27) |
| Liver function test | $2.54 ($0.38) | $2.69 ($0.51) |  | $2.92 ($0.33) | Gamma | (27) |
| Audiometry |  |  |  | $8.80 ($2.33) | Gamma | (27,28) |
| Visual acuity |  |  |  | $7.54 ($1.92) | Gamma | (27) |
| Potassium |  |  |  | $5.37 ($0.81) | Gamma | (27) |
| Biochemistry | $1.79 ($0.24) |  | $32.30 ($4.85) | $2.46 ($1.27) | Gamma | (27) |
| Electrolyte test |  |  |  | $0.95 ($0.14) | Gamma | (27) |
| Bloodgroup RH | $2.54 ($0.38) |  |  | $1.74 ($0.26) | Gamma | (27) |
| Blood clotting | $6.55 ($1.24) |  |  |  | Gamma | (27) |
| Light-emitting diode fluorescence microscopy (LED-FM) | $2.52 ($0.38) |  |  |  | Gamma | (27,28) |
| Magnetic resonance imaging | $2.26 ($0.34) |  |  |  | Gamma | (27) |
| Computerized Tomography (CT) scan | $10.10 ($1.52) |  |  |  | Gamma | (27) |
| Ultrasound test |  | $0.89 ($0.17) | $6.95 ($1.04) |  | Gamma | (27) |
| Other tests | $3.55 ($0.53) |  |  |  |  |  |
| Cost per month for antiretroviral therapy | $23.48 ($3.52) |  |  |  | Gamma | (33) |
| Cost per month for LTFU patients | $3.13 ($0.76) |  |  |  | Gamma | (27,28) |
| Cost per month for end of life state | $3.69 ($0.70) | $1.11 ($0.21) | $12.00 ($2.26) | $1.99 ($0.41) | Gamma | (27,28) |
| Cost per death | $2.42 ($0.45) | $1.44 ($0.27) | $12.00 ($2.26) | $0.42 ($0.08) | n/a | (34) |
| Cost after TB cure | $2.13 ($0.33) | $1.15 ($0.22) | $12.00 ($2.26) | $1.68 ($0.29) | n/a | assumption |
| Cost per month for liver dysfunction | $3.59 ($0.65) | $2.81 ($0.53) |  | $2.05 ($0.36) | Gamma | (27,29) |
| Cost per month for pancreatitis | $38.28 ($3.09) | $18.06 ($3.41) | $49.03 ($3.96) | $26.63 ($2.03) | Gamma | (27,29) |
| Cost per month for anaemia | $1.88 ($0.12) | $0.54 ($0.10) |  |  | Gamma | (27,29) |
| Cost per month for neutropenia |  | $1.70 ($0.32) |  |  | Gamma | (27,29) |
| Cost per month for QTcF prolongation | $1.24 ($0.22) |  |  | $0.70 ($0.12) | Gamma | (27,29) |
| Cost per month for vomiting |  |  | $9.43 ($2.57) |  | Gamma | (27,29) |
| Cost per month for renal disfunction | $0.53 ($0.10) |  |  |  | Gamma | (27,29) |

###### ****Supplementary Table 5 Probabilistic Sensitivity Analysis results****

|  | Mean incremental costs per person | Percent simulations cost saving | Mean DALYs averted per person | Percent simulations averting DALYs |
| --- | --- | --- | --- | --- |
| Philippines |  |  |  |  |
| BPaL | -$245 | 100% | 0.03 | 50% |
| BPaLC | -$152 | 100% | 0.13 | 54% |
| BPaLM | -$199 | 100% | 0.78 | 89% |
| India |  |  |  |  |
| BPaL | -$111 | 99% | 0.01 | 49% |
| BPaLC | -$28 | 74% | 0.09 | 53% |
| BPaLM | -$79 | 94% | 0.70 | 88% |
| South Africa |  |  |  |  |
| BPaL | -$1,171 | 100% | 0.18 | 59% |
| BPaLC | -$1,053 | 100% | 0.26 | 62% |
| BPaLM | -$997 | 100% | 0.83 | 93% |
| Georgia |  |  |  |  |
| BPaL | -$993 | 100% | 0.45 | 71% |
| BPaLC | -$879 | 100% | 0.55 | 78% |
| BPaLM | -$901 | 100% | 1.25 | 97% |
